# Association of Kallikrein Related Peptidase 3 (KLK3) Gene with Dermatophytosis in the UK Biobank cohort

**DOI:** 10.1101/2022.10.09.22280866

**Authors:** Steven Lehrer, Peter H. Rheinstein

## Abstract

In a previous genome wide association study (GWAS) of UK Biobank (UKB) data, we identified one susceptibility locus, Tubulointerstitial Nephritis Antigen (TINAG), with genome wide significance for dermatophytosis. We used genotype calls from file UKB22418. These data are derived directly from Affymetrix DNA microarrays but are missing many genotype calls. Using computationally efficient approaches, UKB has entered imputed genotypes into a second dataset, UKB22828, increasing the number of testable variants by over 100-fold to 96 million variants. In the current study, we used UKB imputed genotypes in UKB22828 to identify dermatophytosis susceptibility loci.

**Methods:** To identify cases of dermatophytosis, we used ICD10 code B35, which covers Tinea barbae, Tinea capitis, Tinea unguium, Tinea manuum, Tinea pedis, Tinea corporis, Tinea imbricata, Tinea cruris, other dermatophytoses, and dermatophytosis, unspecified. We used PLINK, a whole-genome association analysis toolset, to analyze the UKB22828 chromosome files.

**Results:** GWAS summary (Manhattan) plot of the meta-analysis association statistics highlighted two susceptibility loci, TINAG and Kallikrein Related Peptidase 3 (KLK3), with genome wide significance for dermatophytosis.

**Conclusion:** KLK3 may be a dermatophytosis susceptibility gene. KLK3 could affect risk of dermatophytosis, since kallikreins are necessary for normal homeostasis of the skin.

---

Individual factors, including genetics, predispose to dermatophytosis, a common condition. Acute inflammatory forms of dermatophytosis can have no symptoms at all or can be life-threatening. Due to the high cost of therapy, dermatophytosis is a substantial financial burden [1, 2].

In a previous genome wide association study (GWAS) of UK Biobank (UKB) data, we identified one susceptibility locus, Tubulointerstitial Nephritis Antigen (TINAG), with genome wide significance for dermatophytosis [3]. We used genotype calls from file UKB22418. These data are derived directly from Affymetrix DNA microarrays but are missing many genotype calls [4].

The estimation of missing genotype calls using statistical inference is known as genotype imputation. Imputation is increasingly enhancing the number of SNPs accessible in data, not just to fill in gaps left by genotyping errors but also to estimate the genotypes of variants that were not directly tested. Using computationally efficient approaches together with the Haplotype Reference Consortium and UK10K haplotype resources, UKB has entered imputed genotypes into a second dataset, UKB22828, increasing the number of testable variants by over 100-fold to 96 million variants [5].

In the current study, we used UKB imputed genotypes in UKB22828 to identify dermatophytosis susceptibility loci.

## Methods

To identify cases of dermatophytosis, we used ICD10 code B35, which covers Tinea barbae, Tinea capitis, Tinea unguium, Tinea manuum, Tinea pedis, Tinea corporis, Tinea imbricata, Tinea cruris, other dermatophytoses, and dermatophytosis, unspecified.

Data processing was performed on Minerva, a Linux mainframe with Centos 7.6, at the Icahn School of Medicine at Mount Sinai. We used PLINK, a whole-genome association analysis toolset, to analyze the UKB22828 chromosome files [6] and the UK Biobank Data Parser (ukbb parser), a python-based package that allows easy interfacing with the large UK Biobank dataset [7]. We used the R package qqman for the Manhattan and qq plots [8]. Other statistical analyses were done with R and SPSS 26.

We followed quality control procedures [9] that consisted of the following:

1. Missingness of SNPS 0.05: This command excluded SNPs that are missing in a large proportion of the subjects. In this step, SNPs with low genotype calls were removed.
2. Missingness of individuals 0.05: This command excluded individuals who had high rates of genotype missingness. In this step, individuals with low genotype calls were removed.
3. Hardy Weinberg equilibrium 1e-6: This command excluded markers which deviate from Hardy–Weinberg equilibrium.
4. Minor allele frequency (MAF) threshold 0.01: This command included only SNPs above the set MAF threshold.

## Results

We analyzed data from 462,737 subjects. The age at enrollment was 56 ± 8 (mean ± SD). The subjects were 56% women, 44% men, 95% white and British, 15 ± 5 years of education. 532 subjects had dermatophytosis.

GWAS summary (Manhattan) plot of the meta-analysis association statistics, highlighting two susceptibility loci, Kallikrein Related Peptidase 3 (KLK3) and TINAG, with genome wide significance for dermatophytosis, is shown in figure 1. The upper horizontal line indicates the genome wide significance threshold of a P value less than 5×10^×8^.

**Figure 1.**
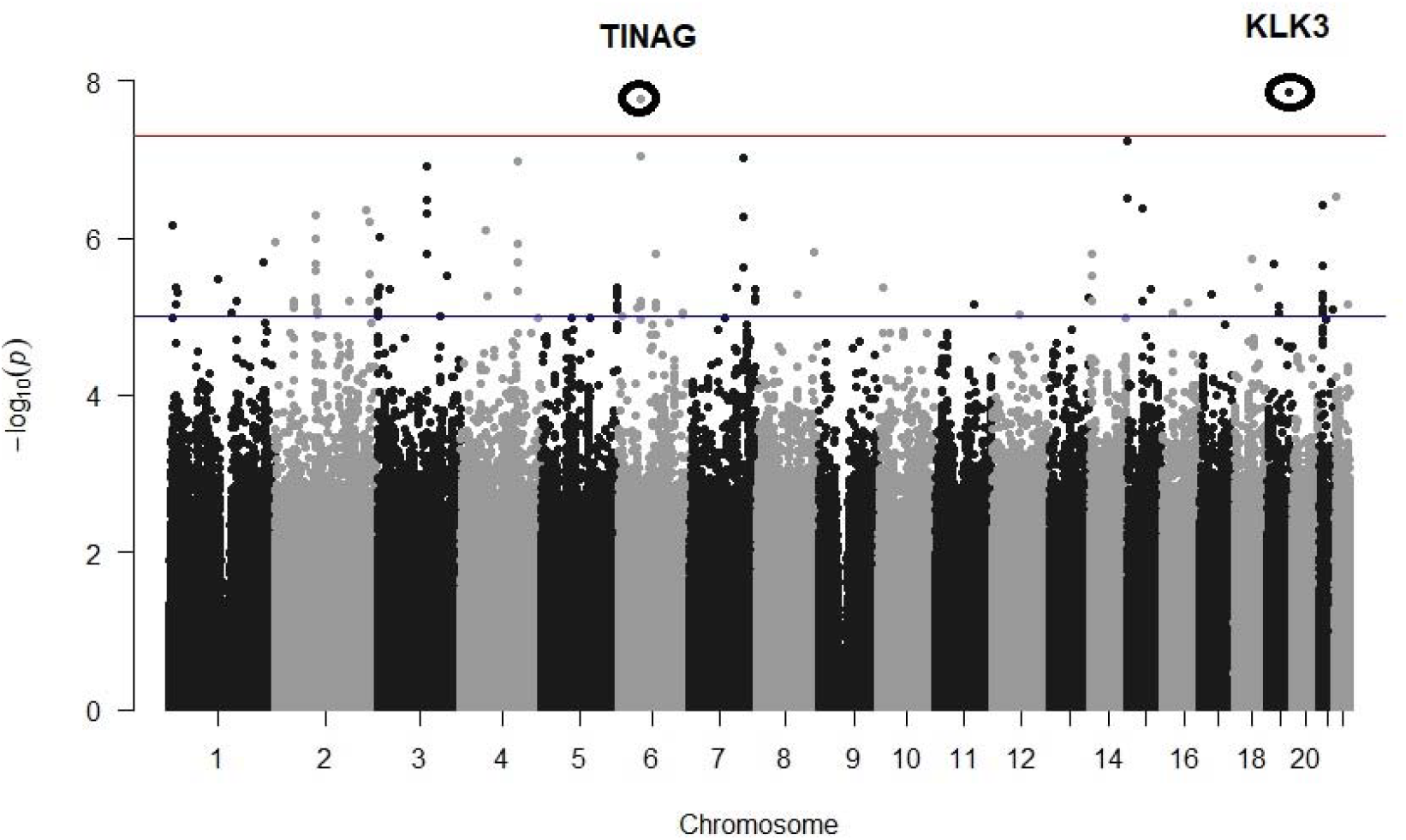
Manhattan plot showing significantly associated dermatophytosis risk loci for TINAG, chromosome 6p12.1, and KLK3, chromosome 19q13.3

The qq plot is shown in figure 2. The x-axis represents expected -log10 (p), y-axis observed - log10 (p) of each SNP. The genomic inflation factor (lambda gc) =1.1. Values up to 1.10 are generally considered acceptable for GWAS and suggest no systematic biases.

**Figure 2.**
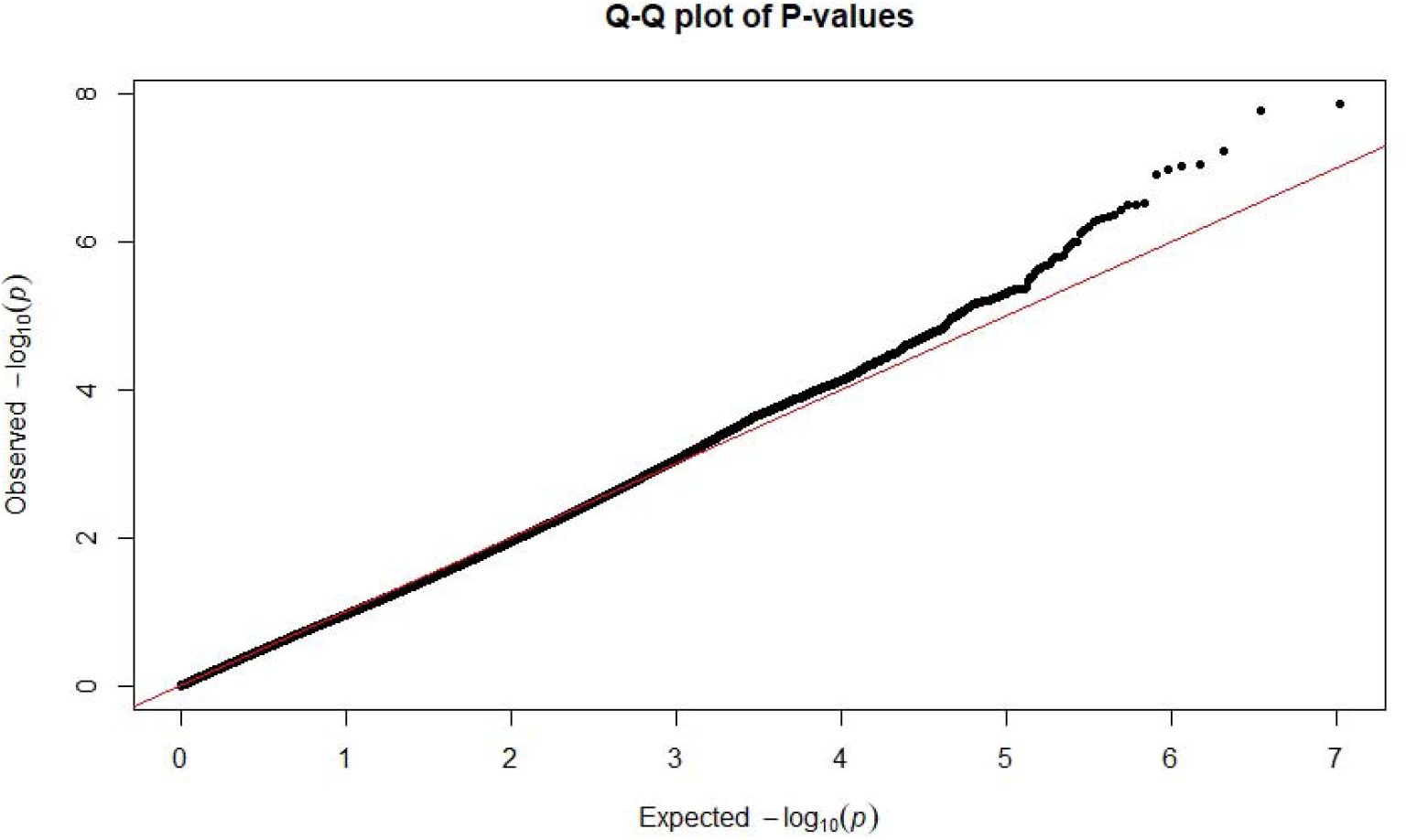
qq plot of p values from GWAS data. Note that most of the p-values observed follow a uniform distribution (left segment of line) but the few that are in linkage disequilibrium with causal polymorphisms produce significant p-values (upper right segment of line). The genomic inflation factor (lambda gc) is 1.1. Values up to 1.1 are generally considered acceptable for GWAS and suggest no systematic biases.

Principal Component Analysis is in figure 3. The x and y axes of variation reduce the data to a small number of dimensions, describing as much variability as possible; they are defined as the top eigenvectors of a covariance matrix between samples [10]. The UKB subjects appear to cluster as a single group, indicating race is not a confounding variable in the GWAS, corroborating the lambda value above. Even though the UK Biobank cohort includes many participants from a variety of ethnic backgrounds, GWAS is possible without sacrificing adequate sample size because most UK Biobank cohort participants report their ethnic background as *British*, within the broader-level group *white*, 88% [11].

**Figure 3.**
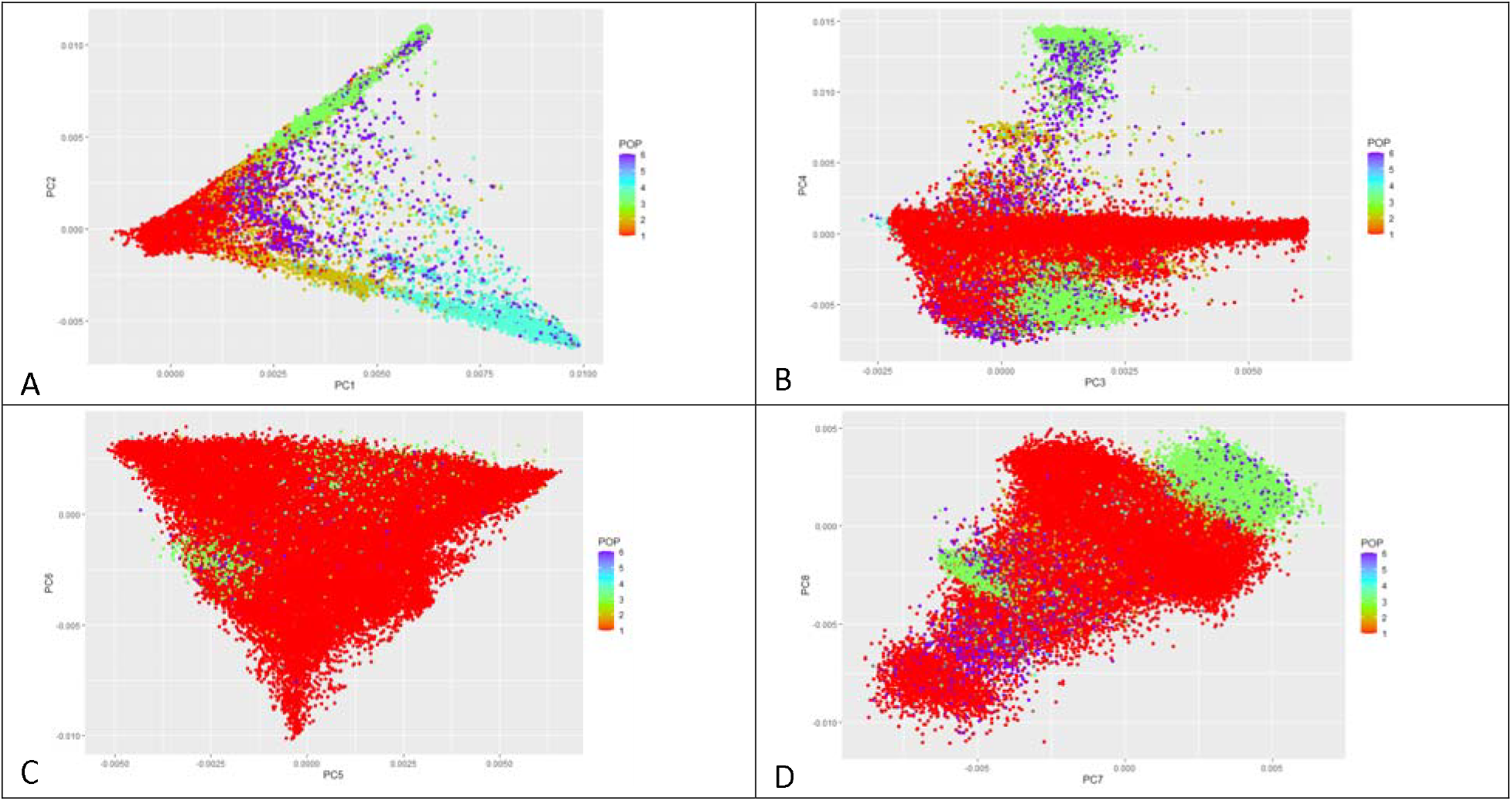
Principal Component (PC) Analysis. POP = population, 1 white, 2 mixed, 3 Asian or Asian British, 4 Black or Black British, 5 Chinese, 6 Other ethnic group. 95% of 487,920 subjects were white British. A) PC1 versus PC2; B) PC3 versus PC4; C) PC5 versus PC6; D) PC7 versus PC8. Principal components represent the directions of the data that explain a maximal amount of variance. Principal Component Analysis tries to put maximum possible information in the first component, then maximum remaining information in the second and so on. The UKB subjects appear to cluster as a single group, especially in B, C, and D, indicating race is not a confounding variable in the GWAS. Even though the UK Biobank cohort includes many participants from a variety of ethnic backgrounds, GWAS is possible without sacrificing adequate sample size because most UK Biobank cohort participants report their ethnic background as *British*, within the broader-level group *white*.

In the GWAS rs61729813 was the SNP most significantly associated with dermatophytosis. rs61729813 is a missense variant within an exon of KLK3, alleles C>G, minor allele frequency (MAF) 0.012. Table 1 contains genotype of rs61729813 versus phenotype, unaffected or mycosis (dermatophytosis) in 462,737 subjects. 0.1% of subjects with genotype CC had dermatophytosis, 0.3% of subjects with genotype CG had dermatophytosis, no subjects were homozygous GG (Fisher exact test two tailed p < 0.001).

**Table 1.** Genotype of rs61729813 versus phenotype, unaffected or mycosis (dermatophytosis) in 462,737 subjects. 0.1% of subjects with genotype CC had dermatophytosis, 0.3% of subjects with genotype CG had dermatophytosis, no subjects were homozygous GG. Fisher exact test two tailed p < 0.001.

| genotype |  | neg | mycosis | Total |
| --- | --- | --- | --- | --- |
| CC | N | 451111 | 503 | 451614 |
|  | % | 99.90% | 0.10% | 100.00% |
| CG | N | 11094 | 29 | 11123 |
|  | % | 99.70% | 0.30% | 100.00% |
| all | N | 462205 | 532 | 462737 |
|  | % | 99.90% | 0.10% | 100.00% |

Results of logistic regression are in Table 2. The dermatophytosis odds ratio (O.R.) for males was 2.19, indicating dermatophytosis is more common in men. The O.R. for age of dermatophytosis was 1.034, in other words dermatophytosis incidence increases with every year of age. Diabetes type 2 increased risk of dermatophytosis, O.R. 2.532. Subjects that were carriers of the G allele of rs61729813 were at increased risk of dermatophytosis (O.R. 2.321).

**Table 2.** Logistic regression with 95% confidence intervals, lower bound (L.B.), upper bound (U.B.). Independent variables sex, age, diabetes type 2, genotype CC versus CG; dermatophytosis present or absent, dependent variable. The dermatophytosis odds ratio (O.R.) for males was 2.19, indicating dermatophytosis is more common in men. The O.R. for age of dermatophytosis is 1.034, in other words dermatophytosis incidence increases with every year of age. Diabetes type 2 increased risk of dermatophytosis, O.R. 2.532. Subjects that were carriers of the G allele of rs61729813 were at increased risk of dermatophytosis (O.R. 2.321).

|  | 95% L.B. | O.R. | 95% U.B. | p value |
| --- | --- | --- | --- | --- |
| Sex | 1.831 | 2.192 | 2.626 | <0.001 |
| Age | 1.021 | 1.034 | 1.046 | <0.001 |
| diabetes type 2 | 2.001 | 2.532 | 3.203 | <0.001 |
| genotype | 1.596 | 2.321 | 3.375 | <0.001 |

## Discussion

KLK3, also known as prostate specific antigen (PSA), belongs to a subclass of serine proteases with a variety of physiological functions. A growing body of research indicates that numerous kallikreins have a role in carcinogenesis and that some of them may serve as innovative cancer and other disease biomarkers. The KLK3 gene is one of 15 members of the kallikrein subfamily that are grouped together on chromosome 19. KLK3 encodes a protease, a single-chain glycoprotein, which is produced in the prostate gland’s epithelial cells and found in seminal plasma. KLK3 protein is believed to hydrolyze high molecular mass seminal vesicle protein during the liquefaction of seminal coagulum. Serum PSA level aids in the detection and follow-up of prostatic cancer. KLK3 gene alternative splicing results in many transcript variants that each encode a different isoform [12].

KLK3 could affect risk of dermatophytosis, since kallikreins are necessary for normal homeostasis of the skin. Keratinocytes of the upper stratum granulosum secrete kallikreins into the stratum corneum. In addition to their function in skin renewal, kallikreins control skin inflammation, the epidermal lipid-rich permeability barrier, and innate immune responses in the epidermis. Kallikreins degrade lipid-processing enzymes and activate the inflammatory response mediator PAR2 on the cell surface of keratinocytes. Antimicrobial cathelicidins, smaller antimicrobial peptides, and pro-inflammatory cytokines all interact with kallikreins. In various skin conditions, including psoriasis, atopic dermatitis, acne rosacea, and Netherton syndrome (NS), kallikrein regulatory function is disrupted [13].

Dermatophytosis is more common in men and diabetics [14, 15]. Our analysis confirms this association and indicates that it is independent of the significant influence of KLK3 (table 2).

A weakness in our study is that single genetic variants and genetic scores composed of multiple variants are associated with birth location within UK Biobank. Major health outcomes appear geographically structured. Haworth et al report that geographic structure in genotype data cannot be accounted for using routine adjustment for study center and principal components derived from genotype data. As a result, coincident structure in health outcomes and genotype data can yield biased associations [16].

We are uncertain why dermatophytosis incidence in the UK Biobank is so low (532/462737). Other sources rank its prevalence in the U.K. as high as 20% [17, 18]. Perhaps most people with dermatophytosis don’t bother to report it to a healthcare professional; or UK Biobank is only capturing patients who are already receiving care for a more serious condition. Indeed, dermatophytosis is associated with diabetes, mentioned above, and end-stage kidney disease [17, 18].

KLK3 may be a dermatophytosis susceptibility gene. More research into genetic and other predisposing factors for dermatophytosis is critical because of the implications for prophylaxis and therapy. It might be possible to prevent infection and recurrence by identifying people who are vulnerable to chronic dermatophytosis. Identifying high-risk families would enable their members to be educated about the dangers of fungal diseases.

## Data Availability

Data in this study may be obtained from UK Biobank after approval of a formal application.

https://www.ukbiobank.ac.uk/

